# Parental Perceptions of Early Childhood Research with In-Home Monitoring: A Qualitative Study

**DOI:** 10.1101/2023.09.11.23295346

**Authors:** Gabriella B. Smith, Mickayla D. Jones, Mary J. Akel, Leonardo Barrera, Marie Heffernan, Patrick C. Seed, Michelle L. Macy, Stephanie A. Fisher, Leena B. Mithal

**Affiliations:** Ann & Robert H. Lurie Children’s Hospital of Chicago and Stanley Manne Children’s Research Institute, Chicago, IL, USA; Department of Pediatrics, Northwestern University Feinberg School of Medicine, Chicago, IL, USA; Division of Maternal-Fetal Medicine, Department of Obstetrics and Gynecology, Northwestern University Feinberg School of Medicine, Chicago, IL, USA

**Author notes:** **Address correspondence to**: Leena B. Mithal, MD MSCI, Division of Pediatric Infectious Diseases, 225 E Chicago Ave, Box #20, Chicago, IL 60611. **Conflict of Interest Disclosures (includes financial disclosures):** The authors have no conflicts of interest relevant to this article to disclose.

## Abstract

**Background:** Despite growing interest in novel approaches to measure home environmental effects on the developing child (the exposome), parental perceptions remain poorly understood. Parents’ perspectives are important for building trust, assessing feasibility, and increasing participation in research that includes home monitoring technologies. We aimed to explore parents’ perceptions regarding these topics.

**Methods:** A diverse group of new and expecting mothers participated in semi-structured interviews. A single interviewer conducted all sessions and introduced a hypothetical longitudinal observational early childhood home-based research study including novel home monitoring approaches: 1) wearable devices, 2) audio monitoring, and 3) environmental sampling. Interviews were audio-recorded, transcribed, and coded. A constant comparative approach identified themes from transcripts.

**Results:** Twenty-four interviews were completed. Emerging themes included 1) Positive Perceptions; 2) Incentives; 3) Transparency and Purpose; 4) Privacy and Safety Concerns; and 5) Logistics and Feasibility. Perceptions were similar across racial, ethnic, and socioeconomic groups. Overall perceptions were positive with several central motivators for hypothetical participation. Participants desired additional information related to feasibility and the purpose of studies. Many had concerns related to wearable device safety and data privacy; a trusting relationship with the research team was a priority.

**Conclusion:** Many participants had positive sentiments regarding longitudinal observational studies involving pregnancy and infancy with various incentives, yet they expressed concerns about safety, privacy, feasibility, and transparency. These findings can inform future perinatal and early childhood research study design, particularly those that novel home monitoring approaches, to ensure protocols and communication are transparent, inclusive, and appealing to parents.

## Introduction

The perinatal and early life period is a critical window for childhood development. ^1–4^ As such, these are important times for researchers to investigate health trajectories and the perinatal origins of disease. ^5^ To effectively study this period, it is important to do so longitudinally and across research domains. ^6^ Most current early childhood research gathers data at clinic visits through surveys and biospecimen collections. ^7,8^ While these data provide valuable information, few studies examine the home environments where children spend most of their early life. ^9,10^ Here, children may be exposed to chemicals, noise, microbes, and stress. ^11–15^ To characterize these pediatric exposomes, researchers must collect data from the household and from children while at home. Innovative methods to study the exposome can capitalize on novel home monitoring approaches, including wearable devices, audio monitoring, and sampling the home environment. ^16–18^

Despite growing interest in perinatal and early childhood research, little is known about parental perspectives regarding study design or novel home monitoring approaches. ^19–21^ There is a gap in knowledge about the perspectives of Black and Hispanic or Latinx families, groups with lower participation in research as a result of historical abuses and exclusion. ^22–26^ As researchers work to increase research participation and ensure positive experiences, understanding the perspectives of new and expecting parents from diverse racial and ethnic groups is crucial. Their sentiments may provide insight on how to build trust and form relationships with families, particularly those skeptical of medical research. ^27^ With more data regarding parental goals, concerns, and questions, researchers will be better equipped to design studies that are novel, inclusive, and appealing to potential participants. In this qualitative investigation, we examined the perceptions of a diverse group of pregnant women and new mothers regarding early childhood research and novel methods for collecting home environmental data.

## Methods

### Recruitment and Participants

Recruitment was conducted in three Obstetrics and Gynecology clinics, two at a large academic center and one at a Federally Qualified Health Center in Chicago, Illinois. Recruitment fliers were distributed, and providers referred potential participants. Recruitment materials were also disseminated in parenting Facebook groups. Interested individuals completed a screening survey with self-reported demographic data, and a purposive sample of English-speaking individuals was invited to participate in a one-hour Zoom interview. Interviews were conducted between September 16, 2022 and January 26, 2023.

The research team invited participants with diverse backgrounds across socioeconomic status (SES), self-reported race/ethnicity, and pregnancy or parenting status. Socioeconomic groups were defined based on annual household income above $50,000 (high SES) or less than or equal to $50,000 (low SES). Represented racial/ethnic groups included non-Hispanic White, non-Hispanic Asian, non-Hispanic Black, or Hispanic/Latina. We prioritized enrolling non-Hispanic Black and Hispanic/Latina participants. As a result, we enrolled comparatively fewer White and Asian participants and grouped these participants together for recruitment and analysis purposes. Participants were either currently pregnant or a new parent, defined as having at least one child under the age of two years. Initially, we aimed to enroll three participants in each of six subcategories (Figure 1). Once this goal was met, interviews were reviewed to determine if thematic saturation had been reached. If thematic saturation was not reached, two more participants were interviewed, and the subgroup was re-reviewed for saturation. This process was repeated until thematic saturation was met by all groups.

**Figure 1.**
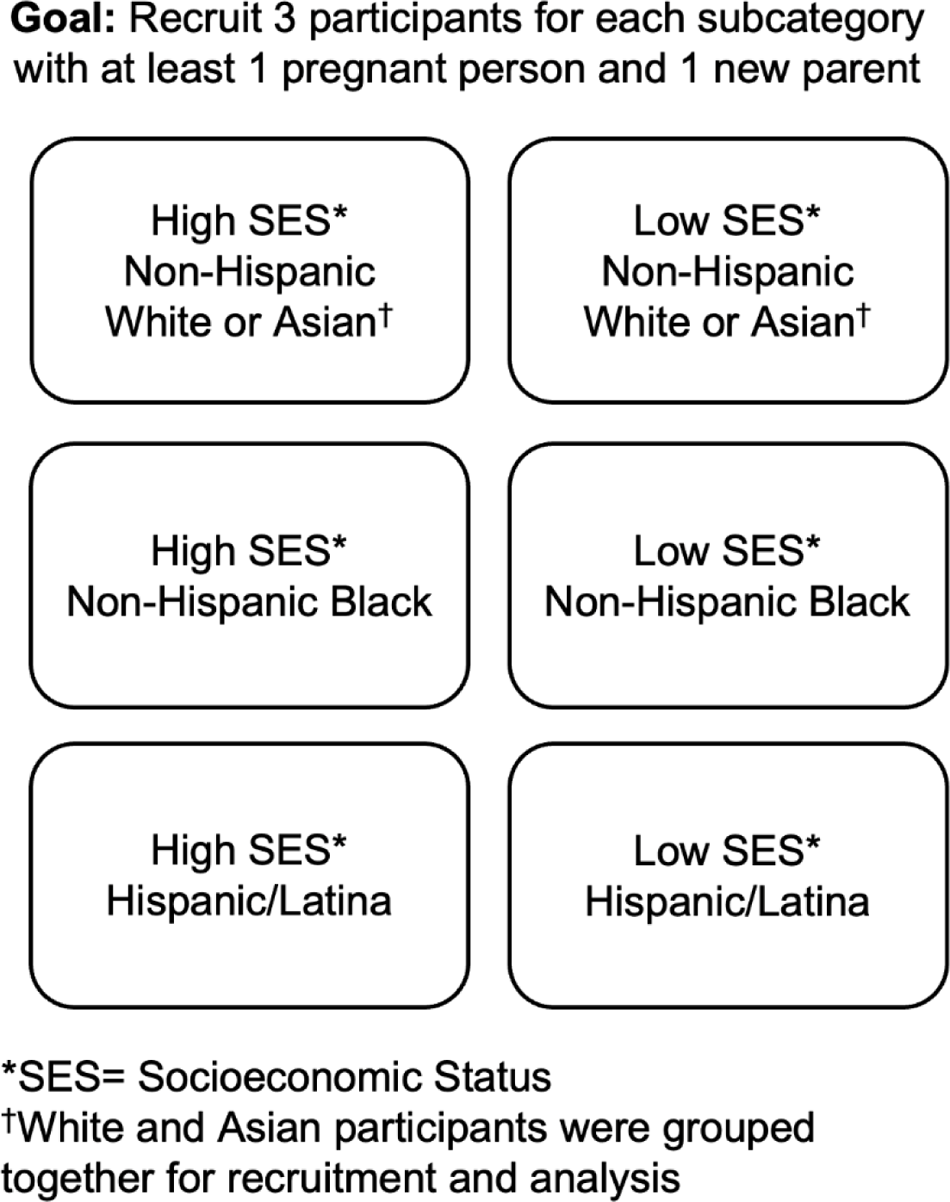
Initial target demographic subcategories to fill during enrollment. After three participants were enrolled in each subcategory, saturation was assessed.

All subjects provided verbal informed consent and were offered a $50 gift card for their participation. This study was approved by the Institutional Review Board of Ann & Robert H. Lurie Children’s Hospital of Chicago (IRB 2022-5534).

### Semi-structured Interviews

One team member (M.J.) conducted all interviews via Zoom using a semi-structured interview guide. The interviewer described four different hypothetical research studies (Table 1). The first was a general longitudinal observational perinatal research study, mirroring the design of an upcoming study at Lurie Children’s and Northwestern Memorial Hospital at the Northwestern University Feinberg School of Medicine. Then, the interviewer described three hypothetical scenarios, each focused on a novel home monitoring approach: 1) wearable devices, 2) audio monitoring, and 3) environmental sample collection. The interviewer probed with follow-up questions about the study structure, modes of data acquisition, logistics, concerns, and incentives.

**Table 1.** Overview of the four hypothetical studies. The descriptions of these studies were adapted from the interview guide.

| Hypothetical Study | Overview |
| --- | --- |
| General Perinatal Research | Researchers want you to participate in a study that would begin in the first months of pregnancy and continue until your baby turns 2 years old. As part of this study, researchers would collect information about your health, your baby's health, and the environment around you both. This information may be collected through surveys, samples from you and your baby, such as blood samples, and samples from your home, such as water samples. |
| Wearable Devices | Researchers want you to participate in a study where you wear a small device on your wrist, like a watch or a sticker, during pregnancy. You would wear a device for a few hours a day each month for a couple months of your pregnancy. Your baby would also wear a device for a few hours a day each month for 3 months after birth. The device would monitor your and your baby's heart rate, motion, location, and body temperature. The device has been tested and found to be safe on pregnant patients and babies. |
| Audio Monitoring | Researchers want to study the relationship between sound and development in early childhood. They want you to participate in a study where there are 1-2 monitors placed in your home that can detect volume and pitch of sound, such as crying, speech, or background noise. |
| Environmental Sample Collection | Researchers want to study the relationship between things in the home environment and your child's health. To participate, a researcher would come to your home a few times and collect samples. The sample could be a small amount of water from your tap or a wipe of the countertops in your home. |

### Analysis

Audio recordings of the Zoom interviews were transcribed, and de-identified transcripts were inductively analyzed by four study team members (M.A., L.B., M.J., G.S.) using Dedoose qualitative software. ^28^

The study team developed an initial codebook using a deductive approach based on pre-existing knowledge and the interview guide. After the initial codebook was developed, each individual member of the coding team coded the same two transcripts. The coding team then met to review their coding for each transcript to establish agreement, ensure reliability, and refine code definitions. Once the codebook was finalized, all subsequent transcripts were coded by a primary and secondary coder who discussed any disagreements. All code excerpts were extracted and reviewed by two team members (L.B., M.J.) to identify themes. A constant comparative approach was used to characterize barriers and facilitators to research participation. The study team met regularly to analyze excerpts and discuss themes.

## Results

### Sample Characteristics

A total of 24 individuals were enrolled and participant demographics reflected the socioeconomic and racial/ethnic diversity we sought (Table 2). All participants identified as female with a median age of 30 years (IQR [27-35]); 46% of participants were currently pregnant. Fifteen (62%) met high SES criteria and 9 (38%) met low SES criteria. One third of participants self-identified as non-Hispanic Black, 33% as Hispanic/Latina, 25% as non-Hispanic White, and 8% as non-Hispanic Asian.

**Table 2.** Demographics of participants. Data are reported as n (%) or median [interquartile range].

|  | **Total**<br>**(n=24)** | **New Parent**<br>**(n=11)** |  | **Currently Pregnant**<br>**(n=13)** |  |
| --- | --- | --- | --- | --- | --- |
|  |  | High SES*<br>(n=7) | Low SES*<br>(n=4) | High SES*<br>(n=8) | Low SES*<br>(n=5) |
| **Age**<br>Median [IQR] | 30 [27-35] | 35 [30-36] | 29 [29-29] | 31 [28-33] | 29 [26-30] |
| **Female Gender**<br>n (%) | 24 (100) | 7 (100) | 4 (100) | 8 (100) | 5 (100) |
| **Race/Ethnicity**<br>n (%) |  |  |  |  |  |
| Non-Hispanic White | 6 (25) | 1 (14) | - | 4 (50) | 1 (20) |
| Non-Hispanic Asian | 2 (8) | 2 (29) | - | - | - |
| Non-Hispanic Black | 8 (33) | 1 (14) | 2 (50) | 3 (38) | 2 (40) |
| Hispanic/Latina | 8 (33) | 3 (43) | 2 (50) | 1 (12) | 2 (40) |

\*High Socioeconomic Status (SES) was defined as annual income greater than \$50,000; low SES was defined as annual income lower than \$50,000

### Themes

Five themes emerged from our interviews: 1) Positive Perceptions; 2) Incentives 3) Transparency and Purpose; 4) Privacy and Safety Concerns; and 5) Logistics and Feasibility (Table 3). There were no stark differences in themes across race, ethnicity, SES, or between pregnant people and new mothers.

**Table 3.** Exemplary quotations for each theme and hypothetical research scenario.

| Exemplary Quotations |  |
| --- | --- |
| **Theme: Positive Perceptions** |  |
| Perinatal Research | “I don't see no red flags, but ... I mean it sounds good. I think it's amazing that they want to know like how it is pregnancy-wise and baby-wise after the baby's born. I totally think that's amazing. I mean I wish when I was younger, because I was a teen mom, so now that I'm older, I wish this was how it was back then, so we like [knew] what's going on ...” *Participant 2023 (Hispanic/Latina, New Parent, Low SES)* |
| Audio Monitoring | “I wouldn't be opposed to that, especially if you can't detect what's being said, but just to be able to detect the pitches of the sound and how loud it is. I think that could be effective to research how babies are brought up and how they could be affected by different tones, voices, and sounds ...” *Participant 2012 (Black, Pregnant, Low SES)* |
| Environmental Sampling | “I think that's good. I think sometimes especially when it comes to the environment. Sometimes you really don't know or think about the things around you that could affect your health, you know...” *Participant 2001 (Black, Pregnant, High SES)* |
| Wearable Devices | “I would say it is a very good idea, because ... you won't miss things out. You'll be able to have a lot of data, you get so much data from it, it's very helpful” *Participant 2013 (White, Pregnant, Low SES)* |
| **Theme: Incentives and Returning Results** |  |
| Perinatal Research | “You could just give one thing at a time. Here goes a box of diapers. Then the following time here goes a box of wipes. You don't always have to give both at once. But that would be perfect ... A box of diapers and wipes? That is not bad at all ... I need that all the time” *Participant 2016 (Hispanic/Latina, New Parent, Low SES)* |
| Audio Monitoring | “... I think it's always nice once some of the data is out to share with the participants 'cause you don't always get that opportunity. Having said that, by the time you get that data, how relevant will it be for that situation? ... But it is always interesting to get that data afterwards.” *Participant 2021 (White, Pregnant, High SES)* |
| Environmental Sampling | “Definitely tell me. Definitely help me research of what I can do to help myself with this problem or my landlord or the person who owns the building. Yes, of course. Definitely give me all the feedback on this one. The same hour or second.” *Participant 2016 (Hispanic/Latina, New Parent, Low SES)* |
| Wearable Devices | “... That would actually be a huge asset if it was a device where I could see the heart rate data for that peace of mind... I actually think that most parents crave data and there's not a lot of it that's reliable about their own kid.” *Participant 2025 (White, New Parent, High SES)* |
| **Theme: Transparency and Purpose** |  |
| Perinatal Research | “Well just coming back to the home visit, one thing that will come up in my mind will be not to trust. Now how can I trust these people coming to my house now? Can I trust them? ... Not just to trust, building trust is very important, because you know if I take it into the house it's something else. So yeah I would like to build that trust relationship.” *Participant 2013 (White, Pregnant, Low SES)*<br>“How is any of this information that I will provide be useful to the researcher and to me? Then I will also want to know if you can be tested and trusted.” *Participant 2019 (Black, Pregnant, High SES)* |
| Audio Monitoring | “I guess like what's the purpose of this study, like the fact that now there's having to be monitors within the home and kind of like invading the home's privacy.” *Participant 2017 (Hispanic/Latina, Pregnant, Low SES)* |
| Environmental Sampling | “Obviously I want the basic information like why are you doing it? What are you collecting? What do you hope to learn from the things that you're collecting in my home.” *Participant 2005 (White, Pregnant, High SES)* |
| Wearable Devices | “Probably why are they doing a study like this, what's behind the study.” *2017 (Hispanic/Latina, Pregnant, Low SES)*<br>“What is this device [collecting] besides the heart rate motion, what data are you trying to get from it? What conclusion are you trying to come to?” *Participant 2017 (Hispanic/Latina, Pregnant, High SES)* |
| **Theme: Privacy and Safety Concerns** |  |
| Perinatal Research | “... I'll also want to know if this information, you earlier said it was going private, but I want to know how private it is because I wouldn't want a situation where my child's life is already out there before he's actually out there.” *Participant 2003 (Black/African American, Pregnant, High SES)* |
| Audio Monitoring | “I'm just scared, what if it gets into the wrong hands? What if I'm on call and I get to disclose things about my workplace, about my family, about myself and then get into the wrong hands? ... After the research what happens? ... I just feel it's kind of invasion of privacy for me.” *Participant 2024 (Black, Pregnant, Low SES)* |
| Environmental Sampling | “I would have to know more about the research because like I said, I'm a very private person, so somebody coming in my home and doing that, I just don't like. I like to be private in my home life.” *Participant 2018 (Hispanic/Latina, Pregnant, Low SES)* |
| Wearable Devices | “You may have some people that may not wanna wear the monitor or be concerned about being tracked with location and stuff...” *Participant 2021 (White, Pregnant, High SES)* |
| **Theme: Logistics and Feasibility** |  |
| Perinatal Research | “If it's something simple that doesn't have a lot of extra time or thought honestly to my life then it's no biggie. But if somebody was coming to my house once a month, and I had to go make special visits for something and something else, then no.” *Participant 2005 (White, Pregnant, High SES)* |
| Audio Monitoring | “One of my questions would be, okay, well, this is the size of my place, do I need one or two or something? Just the logistics of it and how many. I would ask how long would the device be in my home...” *Participant 2002 (Asian, New Parent, High SES)* |
| Environmental Sampling | “As a working parent who works most of the time outside of my home. The first thing is just scheduling this. Oh my gosh, ... they're going to come and sample 15 things in my home. I would want to know like, they would be coming around my availability? They'd be able to come on the weekends and the evenings? Just the logistics of it” *Participant 2025 (White, New Parent High SES)* |
| Wearable Devices | “It's okay but I would need constant reminders because one is going to forget something but probably if there is an app or a kind of reminder that comes up or sort of an alarm like there might be a diary to feel just like that should help” *Participant 2024 (Black, New Parent, Low SES)* |

\*High Socioeconomic Status (SES) was defined as annual income greater than \$50,000; low SES was defined as annual income lower than \$50,000

### Positive Perceptions

Twenty-one (88%) participants expressed positive thoughts regarding the hypothetical general perinatal study. More participants expressed positive sentiments regarding environmental sampling (88%) compared to audio monitoring (79%) or wearable devices (71%). Participants responded more positively to audio monitors that collect the volume and pitch of sound than those that could analyze words (Table 3).

Parents resonated with the sentiment that the perinatal and early childhood period was a formative time in a baby’s life and, therefore, an important time to study:

> “I think that it’s really good because one thing that I’m seeing is that … The early stages of life is really kind of critical for the rest of their lives. It really shapes the type of person that they’re going to be or the type of different triggers that they may have and stuff like that. So I think this is really cool to kind of like do research on something like that”

-Participant 2001 (Black, Pregnant, High SES).

### Incentives

Participants were asked “What would you want to get in return if you participated?” In response, a wide range of incentives emerged (Table 3). “Returning results” was a priority identified in 23 (96%) of interviews, and all 24 participants (100%) wanted environmental sampling data to be returned to them. For several participants, monetary compensation or tangible gifts or resources, such as diapers, wipes, bibs, and toys, would motivate them. Other participants would be motivated by altruism and knowing their involvement could help others. Some cited a combination of these incentives as motivators:

> “For a study like this, I wouldn’t want much. I would just want to know that it’s being used for something very reasonable … I just want to make sure that at the end of the day, at the end of the research, I actually get the data and… I find it very useful actually because I wouldn’t want to put two years into something without getting anything very useful out of it. I would also want to actually just know that okay, this data is going to help a lot of people, families, and I think that will do for me actually.”

-Participant 2003 (Black, Pregnant, High SES)

### Transparency and Purpose

When asked for their opinions, participants wanted to initially understand and be reassured of the purpose of these studies. Mothers also desired transparency surrounding the rationale for study design, the commitment associated with participation, and the potential impact of the research (Table 3).

Parents desired transparency from investigators and emphasized the importance of building trust. Some participants mentioned experiences where they had been skeptical of research in the past:

> “I would just wanna continue to be reassured of the purpose of the study. Yeah, and then just knowing that you guys are honest and trustworthy and that I can trust giving you guys my information, personal information… I think that sometimes these studies can be a little iffy again because you just don’t know what people are doing with the research and if they’re being honest…”

-Participant 2012 (Black, Pregnant, Low SES).

For audio monitoring, participants wanted investigators to be transparent about what exactly was being recorded, how the audio would be shared, and how it would be kept secure. Several parents recommende allowing families to listen to their own study recordings to build trust about what was being recorded.

### Privacy and Safety Concerns

Participants were then asked, “What would your concerns be?” Parents had concerns about the methods for data collection and the security of their personal and health data (Table 3). In our overview of these hypothetical studies, we described measures to ensure security and anonymity of participants’ data beyond the study team, such as deidentification. In response, parents reiterated the importance of confidentiality and assurance that they would not be able to be identified:

> “I would just want to make sure everything is confidential. Like you said, no names and just you can study. I just wouldn’t want anybody to know who I really am …”

-Participant 2017 (Hispanic/Latina, Pregnant, Low SES).

Participants also had specific concerns about the safety and comfort of wearable devices:

> “… How many kids would they [have] tried it on already besides my own? I don’t want my baby [to be] … like a Guinea pig you just pay …”

-Participant 2016 (Hispanic/Latina, New Parent, Low SES).

Concerns emerged about researchers coming into the home to collect samples. Participants would want to know who was coming in and that it was someone they could trust. For audio monitoring, participants worried that the audio data from their homes may not be secure:

> “I don’t like stuff like that. They say that it won’t detect conversation, but it really will I believe and that’s just too weird for someone to … have a lot of information that’s going on in your household. Some things should just be private”

-Participant 2011 (Black, Pregnant, Low SES).

### Logistics and Feasibility

Participants had questions and opinions about study logistics (Table 3). Parents wanted to understand the burden of participation to determine if these studies would be feasible for their families. Fourteen (58%) of participants commented on the feasibility of the general study.

Mothers raised many questions about the time commitment, both how long the study would last and how many visits there would be, especially after delivery. Participants also wanted to know if study visits would take place in person or virtually:

> “I think the big thing is probably time commitment for it. That’s probably the biggest thing with anything is how long, how many times? … And then after I guess what’s the expectation for after [pregnancy] like? Is it on Zoom? Do you have to drive your kid in… ?”

-Participant 2020 (Asian, New Parent, High SES)

Regarding the novel home monitoring approaches, parents had similar logistical questions about coordinating home visits, reminders to put on wearable devices, duration of wearing devices, and the size and number of audio monitoring devices.

## Discussion

This qualitative study examines perspectives among a diverse population of new and expecting mothers on longitudinal observational early childhood research and novel home monitoring approaches. We found participants to have favorable impressions of perinatal research and the potential impact of such studies on their own children’s health and medicine more broadly.

Participants desired a true partnership with the study team, rather than a purely transactional relationship; parents proposed several different examples of acceptable incentives, including wanting to receive relevant results about their child’s health and home environment. At the same time, we observed concerns regarding privacy and safety, a desire for transparency and clarity about the purpose of studies, and questions about logistics and feasibility.

Previous studies have examined parental perspectives regarding research in different pediatric subspecialties, including hospitalized neonates and clinical trials. ^20,26,29–31^ These studies have identified facilitators to research participation, including benefit to their child, altruism, trust, and relationships with research teams, and barriers, such as lack of information, safety concerns, and logistical burdens. ^20,26,29–31^ Few studies investigate the sentiments of new and expecting parents specifically on longitudinal observational research during the perinatal and early childhood period, a uniquely stressful time for many families, or outside of the context of children with active medical issues. ^32^ Even fewer studies seek out the perspectives of Black and Hispanic/Latinx parents as well as families from lower socioeconomic backgrounds. Our study addresses these gaps by identifying the sentiments of a demographically diverse group of mothers regarding both early childhood study design and novel monitoring approaches.

Barriers and facilitators to participating in early childhood research in our study were consistent with prior literature. Snowdon et al. conducted interviews with parents who were approached about their child’s participation in perinatal randomized control trials. While this study provides important insight about parental perceptions and speed of decision making, it focuses on parental views related to the consent process as opposed to study design and engagement. In the Nathe et al. systematic review of parental views of various pediatric research studies, facilitators to research included the importance of research, altruism, and health benefit to child. ^20^ Barriers, such as burden of participation, risk to child, and lack of trust, also emerged. ^20^ To build on the existing literature, we were able to analyze the interactions between different facilitators and barriers. Additionally, we present ideas from parents on how to address concerns and amplify motivators.

Despite existing disparities in research participation, ^23,26^ we saw no stark differences in the responses of our participants across race, ethnicity, or SES or between pregnant people and new parents. Our findings may be considered when working to build partnerships and trust with families, including those from communities that have faced racism, mistreatment, and abuse in medicine and research. ^22–26^ Previous studies have determined factors that impact parental willingness to consent to research, including lack of trust and logistical challenges. ^29–31^ Our study identifies practices that address these factors and, therefore, enhance opportunities to achieve equitable parental participation.

Building trust with participants and being transparent both during recruitment and throughout the study duration should be a top priority for investigators. Researchers should aim to align the incentives of their studies with the motivations of parents and families, and there should be an effort to include multiple types of incentives, including monetary compensation, resources, returning results, and highlighting the altruistic impact of participation in longitudinal observational research studies. Specifically, study teams may consider providing regular updates and returning results at study visits or upon parental request.

Our results indicate that researchers need to balance their desire for robust data with the burden of participation for families. Researchers could achieve this balance by partnering with parents to design logistically practical studies that accommodate different families’ schedules. Due to the COVID-19 pandemic, remote study visits have emerged as one such way to provide additional convenience in some cases. ^33^ Logistical details, such as time commitment and location of visits, should be presented clearly when approaching potential participants. In this spirit of transparency, it is also important to discuss safety and privacy concerns directly with families and to be clear about the rationale underlying study design, what data is collected, how it is kept secure, and the risk of potential data breaches.

The insights from this study should be considered when designing future early childhood research studies implementing novel home monitoring approaches. When investigators are designing future longitudinal, time-intensive studies, their success may be increased by conducting qualitative research as a prelude to gauge parental perspectives before finalizing protocols and launching enrollment. Study designs that engage families in ongoing feedback may also promote successful recruitment. With increased insight into parental concerns and sentiments, investigators can work to design studies that will enhance the experiences of families, increase longitudinal research engagement, and improve retention of participants.

There are several limitations to this study. Due to the qualitative nature of this study, our findings may not be widely generalizable. We analyzed the sentiments of individuals who were willing to participate in our study, and, therefore, participation bias limits our ability to understand perspectives of parents who are not interested in participating in any type of research. We were able to recruit a diverse sample, but all participants were female and spoke English. Future studies should aim to engage fathers, non-English speaking individuals, and those who have limited interest in participation in studies. Additionally, larger survey studies about parental perspectives may yield results that can be generalized more broadly to inform study design and best practices. Finally, we asked participants to share their perspectives about hypothetical studies, which may differ from how parents would feel about an actual longitudinal observational study opportunity.

## Conclusion

New and expecting mothers view research during early childhood and novel home monitoring methods approaches positively but also have some concerns that should be considered in future study design. Parents conveyed a desire for researchers to build trusting partnerships and reduce the burdens of participation. Further qualitative studies are warranted to explore additional perspectives of fathers, linguistically diverse parents, and parents who are more cautious about research participation. These qualitative findings can contribute to larger mixed-methods studies to yield further generalizable results. Investigators can leverage the perceptions of parents identified in our study to inform future design and implementation of early childhood studies of the exposome, which may increase rates and satisfaction of participation.

## Data Availability

All data produced in the present study are available upon reasonable request to the authors.

## Acknowledgements

We would like to thank the Lurie Children’s Founders’ Board for their funding support for this study. We would like to acknowledge Erie Family Health Center and Prentice Ambulatory Clinic for supporting our recruitment efforts in their clinics. This work was conducted with support from faculty and staff in the Mary Ann & J. Milburn Smith Child Health Outcomes, Research, and Evaluation Center’s Catalyst at Stanley Manne Children’s Research Institute at Ann & Robert H. Lurie Children’s Hospital of Chicago.

## Abbreviations

SES: Socioeconomic Status

## Notes

**Funding/Support**: All phases of this study were supported by the Founders’ Board of Lurie Children’s Hospital.

### Competing Interest Statement

The authors have declared no competing interest.

### Funding Statement

All phases of this study were supported by the Founders' Board of Lurie Children's Hospital.

### Author Declarations

The IRB of Ann & Robert H. Lurie Children's Hospital gave ethical approval for this work (IRB 2022-5534).

## References

1. Arima Y, Fukuoka H. Developmental origins of health and disease theory in cardiology. J Cardiol. Jul 2020;76(1):14–17. doi:10.1016/j.jjcc.2020.02.003

2. Kalbermatter C, Fernandez Trigo N, Christensen S, Ganal-Vonarburg SC. Maternal Microbiota, Early Life Colonization and Breast Milk Drive Immune Development in the Newborn. Front Immunol. 2021;12:683022. doi:10.3389/fimmu.2021.683022

3. Nobile S, Di Sipio Morgia C, Vento G. Perinatal Origins of Adult Disease and Opportunities for Health Promotion: A Narrative Review. J Pers Med. Jan 25 2022;12(2)doi:10.3390/jpm12020157

4. Simeoni U, Armengaud JB, Siddeek B, Tolsa JF. Perinatal Origins of Adult Disease. Neonatology. 2018;113(4):393–399. doi:10.1159/000487618

5. Barker DJ. The origins of the developmental origins theory. J Intern Med. May 2007;261(5):412–7. doi:10.1111/j.1365-2796.2007.01809.x

6. Paneth N, Monk C. The importance of cohort research starting early in life to understanding child health. Curr Opin Pediatr. Apr 2018;30(2):292–296. doi:10.1097/mop.0000000000000596

7. Magnus P, Birke C, Vejrup K, et al. Cohort Profile Update: The Norwegian Mother and Child Cohort Study (MoBa). Int J Epidemiol. Apr 2016;45(2):382–8. doi:10.1093/ije/dyw029

8. Olsen J, Melbye M, Olsen SF, et al. The Danish National Birth Cohort--its background, structure and aim. Scand J Public Health. Dec 2001;29(4):300–7. doi:10.1177/14034948010290040201

9. Faro EZ, Sauder KA, Anderson AL, et al. Characteristics of Environmental influences on Child Health Outcomes (ECHO) Cohorts Recruited During Pregnancy. MCN Am J Matern Child Nurs. Jul-Aug 01 2021;46(4):230–235. doi:10.1097/nmc.0000000000000725

10. Ferguson KT, Cassells RC, MacAllister JW, Evans GW. The physical environment and child development: an international review. Int J Psychol. 2013;48(4):437–68. doi:10.1080/00207594.2013.804190

11. Lu C, Liu Z, Liao H, Yang W, Li Q, Liu Q. Effects of early life exposure to home environmental factors on childhood allergic rhinitis: Modifications by outdoor air pollution and temperature. Ecotoxicol Environ Saf. Oct 1 2022;244:114076. doi:10.1016/j.ecoenv.2022.114076

12. Lu Y, Lin S, Lawrence WR, Lin Z, Gurzau E, Csobod E, Neamtiu IA. Evidence from SINPHONIE project: Impact of home environmental exposures on respiratory health among school-age children in Romania. Sci Total Environ. Apr 15 2018;621:75–84. doi:10.1016/j.scitotenv.2017.11.157

13. Norbäck D, Zhang X, Tian L, et al. Prenatal and perinatal home environment and reported onset of wheeze, rhinitis and eczema symptoms in preschool children in Northern China. Sci Total Environ. Jun 20 2021;774:145700. doi:10.1016/j.scitotenv.2021.145700

14. Vardoulakis S, Giagloglou E, Steinle S, et al. Indoor Exposure to Selected Air Pollutants in the Home Environment: A Systematic Review. Int J Environ Res Public Health. Dec 2 2020;17(23)doi:10.3390/ijerph17238972

15. Wang J, Norbäck D. Home environment and noise disturbance in a national sample of multi-family buildings in Sweden-associations with medical symptoms. BMC Public Health. Nov 3 2021;21(1):1989. doi:10.1186/s12889-021-12069-w

16. Istrate D, Binet M, Cheng S. Real time sound analysis for medical remote monitoring. Annu Int Conf IEEE Eng Med Biol Soc. 2008;2008:4640–3. doi:10.1109/iembs.2008.4650247

17. Lytle DA, Formal C, Cahalan K, Muhlen C, Triantafyllidou S. The impact of sampling approach and daily water usage on lead levels measured at the tap. Water Res. Jun 1 2021;197:117071. doi:10.1016/j.watres.2021.117071

18. Sharma A, Badea M, Tiwari S, Marty JL. Wearable Biosensors: An Alternative and Practical Approach in Healthcare and Disease Monitoring. Molecules. Feb 1 2021;26(3)doi:10.3390/molecules26030748

19. Hubbell BJ, Kaufman A, Rivers L, et al. Understanding social and behavioral drivers and impacts of air quality sensor use. Sci Total Environ. Apr 15 2018;621:886–894. doi:10.1016/j.scitotenv.2017.11.275

20. Nathe JM, Oskoui TT, Weiss EM. Parental Views of Facilitators and Barriers to Research Participation: Systematic Review. Pediatrics. Jan 1 2023;151(1)doi:10.1542/peds.2022-058067

21. Schramm K, Grassl N, Nees J, et al. Women’s Attitudes Toward Self-Monitoring of Their Pregnancy Using Noninvasive Electronic Devices: Cross-Sectional Multicenter Study. JMIR Mhealth Uhealth. Jan 7 2019;7(1):e11458. doi:10.2196/11458

22. López-Cevallos DF, Harvey SM, Warren JT. Medical mistrust, perceived discrimination, and satisfaction with health care among young-adult rural latinos. J Rural Health. Fall 2014;30(4):344–51. doi:10.1111/jrh.12063

23. Rees CA, Stewart AM, Mehta S, et al. Reporting of Participant Race and Ethnicity in Published US Pediatric Clinical Trials From 2011 to 2020. JAMA Pediatr. May 1 2022;176(5):e220142. doi:10.1001/jamapediatrics.2022.0142

24. Scharff DP, Mathews KJ, Jackson P, Hoffsuemmer J, Martin E, Edwards D. More than Tuskegee: understanding mistrust about research participation. J Health Care Poor Underserved. Aug 2010;21(3):879–97. doi:10.1353/hpu.0.0323

25. Shaw MG, Morrell DS, Corbie-Smith GM, Goldsmith LA. Perceptions of pediatric clinical research among African American and Caucasian parents. J Natl Med Assoc. Sep 2009;101(9):900–7. doi:10.1016/s0027-9684(15)31037-3

26. Weiss EM, Olszewski AE, Guttmann KF, et al. Parental Factors Associated With the Decision to Participate in a Neonatal Clinical Trial. JAMA Netw Open. Jan 4 2021;4(1):e2032106. doi:10.1001/jamanetworkopen.2020.32106

27. Webb Hooper M, Mitchell C, Marshall VJ, et al. Understanding Multilevel Factors Related to Urban Community Trust in Healthcare and Research. Int J Environ Res Public Health. Sep 6 2019;16(18)doi:10.3390/ijerph16183280

28. Dedoose version 8.3.17. SocioCultural Research Consultants, LLC. http://www.dedoose.com

29. Hoehn KS, Wernovsky G, Rychik J, Gaynor JW, Spray TL, Feudtner C, Nelson RM. What factors are important to parents making decisions about neonatal research? Arch Dis Child Fetal Neonatal Ed. May 2005;90(3):F267–9. doi:10.1136/adc.2004.065078

30. Kick K, Assfalg R, Aydin S, et al. Recruiting young pre-symptomatic children for a clinical trial in type 1 diabetes: Insights from the Fr1da insulin intervention study. Contemp Clin Trials Commun. Sep 2018;11:170–173. doi:10.1016/j.conctc.2018.08.004

31. Mazzocco MMM, Myers GF, Harum KH, Reiss AL. Children’s Participation in Genetic Prevalence Research: Influences on Enrollment and Reports of Parent Satisfaction1. Journal of Applied Social Psychology. 1999;29(11):2308–2327. 10.1111/j.1559-1816.1999.tb00112.x

32. Snowdon C, Elbourne D, Garcia J. “It was a snap decision”: parental and professional perspectives on the speed of decisions about participation in perinatal randomised controlled trials. Soc Sci Med. May 2006;62(9):2279–90. doi:10.1016/j.socscimed.2005.10.008

33. Kissler K, Breman RB, Carlson N, Tilden E, Erickson E, Phillippi J. Innovations in Prospective Perinatal Research as a Result Of the COVID-19 Pandemic. J Midwifery Womens Health. Mar 2022;67(2):264–269. doi:10.1111/jmwh.13329

